# Deployment-readiness audit of calibration, clinical utility, and fairness in perioperative infection prediction

**DOI:** 10.64898/2026.06.15.26355656

**Authors:** Hugo Guillen-Ramirez, Katharina Lucia Lucas, Yves Max Wintsch, Tobias Ueli Blatter, Karen Triep, Olga Endrich, Guido Beldi

## Abstract

**Objective:** Clinical risk scores intended to guide patient-level decisions can show strong average performance. However, predicted probabilities can be systematically too high or too low in specific subgroups even when overall performance is strong. We audited deployment readiness of a strong end-of-surgery postoperative infection model across clinically relevant subgroups and tested mitigation strategies in miscalibrated subgroups.

**Materials and Methods:** We analyzed out-of-fold predictions for 10,719 surgical procedures at a Swiss tertiary hospital, with 504 postoperative bacterial infection events. Prespecified axes were recorded sex, age stratum, and an EHR-derived physiological-reserve proxy. Within subgroups and pairwise intersections, we evaluated discrimination, calibration, threshold-specific errors, and decision-curve net benefit at the prespecified operating threshold. We compared group-specific isotonic recalibration with Wasserstein-barycenter postprocessing and demonstrated portability in SUPPORT2.

**Results:** Overall AUROC was 0.876. While sex-marginal discrimination was similar in women and men (0.878 vs 0.875), age and reserve stratification revealed deployment-readiness failures. Calibration-in-the-large ranged from -0.86 in frail patients to -2.47 in non-frail patients. At the 0.10 operating threshold, decision-curve net benefit was positive in frail patients but negative in pre-frail and non-frail patients. Isotonic recalibration corrected average physiological-reserve-stratified calibration without worsening Brier scores, whereas Wasserstein postprocessing worsened calibration in most procedure clusters.

**Discussion:** Discrimination-only or sex-marginal evaluation would have missed subgroup failures with clinical-utility implications.

**Conclusion:** Subgroup fairness audits for clinical deployment should jointly evaluate discrimination, calibration, and utility. We implemented the audit as the open-source *isitfair* framework for identifying deployment-relevant subgroup failures, comparing mitigation strategies, and generating structured reports.

## BACKGROUND

Postoperative bacterial infection drives preventable morbidity, length of stay, cost, and mortality [1–3]. Although infections are typically apparent by postoperative day 5 and C-reactive protein can shift identification to about day 3 [4], our prior work aimed to identify risk as early as the end of surgery. We first used 91,794 perioperative EHR records to predict infection within two days (AUROC 0.83), with strategic missing-laboratory imputation improving recall [5]. We then used interpretable summary, trend, and distributional descriptors of intraoperative vital signs in 10,719 procedures, achieving AUROC 0.88 (95% CI, 0.85-0.91), cluster-level calibration, and SHAP-based interpretability [6]. However, strong aggregate discrimination is not sufficient evidence of subgroup deployment readiness.

Strong average performance can mask subgroup degradation. Examples in medical AI are severity underestimation in Black patients [7], underdiagnosis by chest-radiograph classifiers in women and younger patients [8], and temporal fairness drift in postsurgical risk scores despite maintained aggregate calibration [9]. Accuracy and calibration can degrade, and fairness gaps increase, even while aggregate discrimination remains stable [9]. Potential causes for such subgroup performance differences include unrepresentative training cohorts, unmodeled sex-associated differences [10], incomplete inclusion of physiological reserve [11], varying signal quality [12,13], and differences in surveillance intensity or data completeness [14–20].

Accordingly, TRIPOD+AI recommends reporting model performance, including uncertainty, within clinically relevant sociodemographic and clinical subgroups [21]. Fairness and deployment readiness in clinical AI is therefore a joint assessment of discrimination, calibration, and clinical utility, not error-rate parity alone [22,23]. However, workflows for tabular or time-series clinical models remain rare [24]. Current toolkits either target medical imaging [25,26] or are general-purpose tools [27,28], leaving limited support for calibration-focused subgroup fairness assessment and clinical-utility reporting for deployment readiness in clinical data.

We retrospectively audited a previously developed end-of-surgery infection prediction model across recorded sex, age stratum, and a three-level EHR-derived physiological-reserve classification. For each axis, we evaluated three clinically distinct dimensions: (i) discrimination, whether infected patients received higher predicted risks than non-infected patients; (ii) calibration, whether predicted probabilities matched observed risks; and (iii) clinical utility, whether acting at the intended threshold provided net benefit. We implemented this workflow as *isitfair*, an open-source Python package for subgroup fairness auditing, mitigation comparison, and structured reporting of clinical prediction models.

## MATERIALS AND METHODS

### Cohort, model, and outcome

The audited cohort comprised 10,719 surgical procedures performed at Inselspital, Bern University Hospital between 2014 and 2022, including 504 postoperative bacterial infection events. The source cohort was not restricted to operating-room or invasive cases (Supplementary Table 1). The cohort, outcome definition, model architecture, and training pipeline were those of the previously reported end-of-surgery model [6]. Briefly, the model used summary, trend, and distributional descriptors from intraoperative vital-sign time series to estimate the probability of postoperative bacterial infection at the end of surgery. We audited out-of-fold predictions from 5-fold stratified cross-validation with the Synthetic Minority Oversampling Technique (SMOTE) applied within each training fold, so each procedure received a prediction from a model that had not seen that procedure during training.

The outcome was bacterial postoperative infection during the index admission, defined as the continuous inpatient stay containing the surgery, from admission to discharge, using the prespecified ICD-10-GM coding pipeline from the source cohort. Present-on-admission flags, where available, were used to exclude infections coded as already present at admission.

### Sensitive attributes and subgroup axes

The audit prespecified three subgroup axes available at surgery: recorded sex, age group, and physiological reserve. Recorded sex and age were sociodemographic audit axes, whereas physiological reserve was clinical. Physiological reserve was included because unnecessary monitoring escalation was expected to have different consequences by baseline reserve. Recorded sex was extracted from the administrative hospital information system and was binary in the source data. Gender identity was not captured; analyses therefore refer to recorded sex, not gender. Age was grouped as 0-17, 18-44, 45-64, 65-79, and 80 years or older. These bins do not map exactly to the institutional pediatric pathway, which covers patients aged 15 years or younger, or to standard adolescent and young-adult definitions; the 0-17 stratum therefore combines young children and adolescents.

The physiological-reserve proxy was a three-level variable (frail, pre-frail, non-frail) combining the American Society of Anesthesiologists (ASA) score, Hospital Frailty Risk Score (HFRS) [11], and a four-item modified Frailty Index (mFI-4; mFI-5 [29] without functional status, unavailable in our EHR). Components were not analyzed separately. Frail required ASA-PS ≥4, HFRS >15, or mFI-4 ≥2; pre-frail required no frail criterion but ASA-PS 3, HFRS 5-15, or mFI-4 = 1; all remaining patients were non-frail. The mFI-4 comprised diabetes mellitus, chronic obstructive pulmonary disease, congestive heart failure, and hypertension; item definitions and HFRS weights are provided in Supplementary Tables 2 and 3. Because hypertension was approximated from ICD-10 codes without medication linkage, we also evaluated an ASA/HFRS-only reserve definition (Supplementary Table 4). We use “frail,” “pre-frail,” and “non-frail” only as labels for the reserve proxy, not as diagnoses of a validated frailty syndrome. Surgical specialty and procedure cluster were case-mix reference axes. Pairwise intersections were prespecified for sex by age, sex by physiological reserve, and age by physiological reserve.

### Fairness metrics

We evaluated subgroup performance across discrimination, calibration, and clinical utility: whether infected patients received higher scores, whether predicted risks matched observed risks, and whether flagging patients for intensified surveillance, here termed monitoring, at the operating threshold did more good than harm.

#### Discrimination

We assessed whether patients who developed infection received higher predicted risks using AUROC and AUPRC. We prespecified an operating threshold of 0.10: a predicted risk ≥10%, more than double the 4.7% baseline, triggered intensified monitoring, not invasive intervention [30]. Monitoring denotes closer surveillance, such as more frequent vital-sign and laboratory checks and earlier clinical review. The low threshold corresponds to treating a missed infection as approximately nine times as costly as one unnecessary monitoring escalation [30,31]. We also reported sensitivity, specificity, and threshold-specific error-rate gaps, namely the equal-opportunity difference (sensitivity gap), the predictive-equality difference (false-positive-rate gap), the equalized-odds gap (the larger of the sensitivity and false-positive-rate gaps), and the statistical parity difference (difference in the proportion of patients flagged) at this threshold [32,33].

#### Calibration

Calibration assesses whether predicted probabilities correspond to observed risks, for example, whether patients assigned a 10% risk develop infection about 10% of the time. We summarized it with calibration curves, calibration-in-the-large (the intercept), calibration slope, Brier score, expected calibration error, and the integrated calibration index [34–37]. Because overall calibration can conceal subgroup bias, each measure was computed within every subgroup.

#### Clinical utility

Clinical utility assesses whether acting on the score improves decisions at a chosen threshold. We assessed clinical utility using decision-curve net benefit [38] across thresholds from 0.02 to 0.40 and at the prespecified 0.10 operating threshold. Because net benefit depends on outcome prevalence, standardized net benefit was also reported [23]. Full metric definitions and estimator equations are provided in the Supplementary Methods.

### Calibration within subgroups

Subgroup calibration was prespecified because overall calibration may conceal clinically relevant over-or underprediction. Each cohort-level calibration measure was estimated within every subgroup because deployment uses individual probabilities, not only ranks. Overestimation can trigger unnecessary monitoring, delayed discharge, or avoidable antibiotics; underestimation can miss patients needing surveillance. We assessed calibration visually, parametrically, and with aggregate scores.

#### Visual calibration curves

For each subgroup, we plotted the observed infection rate against predicted risk using LOESS smoothing, with 95% bootstrap bands. The 45° identity line represents perfect calibration; a curve below it indicates that the model overestimates risk in that subgroup, whereas a curve above it indicates underestimation.

#### Parametric summaries

We estimated calibration intercept and slope on the log-odds scale [34]. The intercept (calibration-in-the-large) measures systematic over- or underprediction: zero is unbiased; a negative value indicates that the model overestimates risk on average. The slope measures whether the spread of predicted risks is correctly scaled: one is ideal; below one indicates predictions that are too extreme, above one indicates predictions that are too compressed.

#### Aggregate scores

We calculated the Brier score, expected calibration error (ECE; mean absolute predicted-observed gap across ten equal-frequency bins), and integrated calibration index (ICI; the same gap evaluated patient-wise against the LOESS curve). Hosmer-Lemeshow tests were omitted because they are sample-size dependent [39]. Estimator equations are given in the Supplementary Methods.

### Clinical utility within subgroups

Clinical utility was prespecified because calibration alone does not show whether acting on the score improves decisions. We used subgroup decision-curve analysis across t = 0.02 to 0.40. Net benefit counts true positives and subtracts false positives, weighted by the threshold-implied trade-off; positive values improve over monitoring no one, and usefulness also requires comparison with monitor-all. Because net benefit depends on prevalence, we also report standardized net benefit, where 1 is a hypothetical perfect model and 0 is no better than monitoring no one. The primary clinical-utility gap was the maximum absolute pairwise subgroup difference in net benefit across thresholds, with bootstrap CIs. Threshold-specific results were re-expressed as natural frequencies per 1,000 patients.

### Bias-mitigation strategies

Two prespecified post hoc score transformations that do not require model retraining were compared. Group-specific isotonic recalibration [40] was fitted within recorded sex, physiological reserve, and procedure-cluster strata. To avoid in-sample evaluation of mitigated probabilities, both postprocessing strategies were fitted with 5-fold cross-fitting: each transformation was estimated on four folds and applied to the held-out fold, yielding one out-of-fold mitigated prediction per procedure. Wasserstein-barycenter postprocessing [41–43] was fitted on the procedure-cluster axis as a score-distribution mitigation benchmark, because distributional shift was most pronounced on this case-mix axis. The Wasserstein transformation was constrained to demographic parity and treated as a parity-focused comparator rather than a calibration-preserving mitigation [44]. Mitigations were evaluated by their effect on discrimination, calibration, Brier score, and subgroup gaps. Mitigation was prespecified as favorable only when it improved the affected subgroup or corrected calibration without degrading absolute subgroup performance. Because isotonic regression can overfit sparse strata, we also inspected post-mitigation calibration slope and subgroup event counts.

### Portability analysis

To assess portability beyond perioperative prediction, we applied the workflow to SUPPORT2 [45] using a logistic-regression model for in-hospital mortality. This prespecified analysis repeated the Bern reporting sequence: subgroup discrimination, calibration, threshold-specific performance, and clinical-utility summaries. SUPPORT2 was not external validation of the infection model; it was a public-dataset workflow demonstration and allowed evaluation of a race/ethnicity axis unavailable in the Swiss EHR.

### Statistical analysis and reporting

Confidence intervals were estimated using 200 stratified bootstrap resamples. Gap statistics were recomputed within each bootstrap sample to preserve dependence between subgroup estimates. For sparse intersectional cells with fewer than 50 infection events, empirical-Bayes shrinkage was applied toward the relevant parent subgroup; cell counts, event counts, and shrinkage factors are reported in the Supplementary Materials. Multiple testing was controlled using Benjamini-Hochberg false discovery rate correction within prespecified test families. Prespecified sex by physiological reserve, sex by ASA, and sex by comorbidity interaction tests were performed as secondary explanatory analyses and are reported in the Supplementary Materials.

Analyses were implemented in Python using the *isitfair* package developed for this study. The manuscript and supplement follow TRIPOD+AI reporting principles for subgroup performance, calibration, and intended use. Code, package versions, random seeds, and intermediate subgroup outputs are described in the Supplementary Materials.

## RESULTS

### Cohort and subgroup distribution

The audited cohort (Table 1) comprised 10,719 surgical procedures with 504 postoperative infection events (4.7%). The recorded sex was 34.4% female (n = 3,689) and 65.6% male (n = 7,030). Physiological reserve classified 6,139 patients (57.3%) as frail, 3,558 (33.2%) as pre-frail, and 1,022 (9.5%) as non-frail. Infection prevalence increased monotonically with reserve loss (0.6%, 2.8%, and 6.5%, respectively). Baseline differences by recorded sex suggested sex-associated case mix and patient selection rather than direct infection-biology differences. For example, cardiac procedures accounted for 41.7% of operations in men versus 22.7% in women (Cramér’s V = 0.31); men were more often classified as frail (63.6% vs 45.2%) and ASA 4 (57.5% vs 38.8%), whereas women were over-represented at both age extremes (17.1% aged 0-17 or 80+ vs 10.6% of men). Thus, sex-marginal results needed interpretation alongside age, reserve, and procedure mix.

**Table 1.** Cohort characterization by sex. Counts (column percentages) for categorical variables and median [Q1, Q3] for continuous variables, stratified by administratively recorded sex. p-values are from the χ² test for categorical variables and the Mann–Whitney U test for continuous variables. FDR is the Benjamini–Hochberg-adjusted q-value across the eight tested variables, controlling the false discovery rate at q < 0.05. Physiological reserve is the three-level composite of ASA Physical Status, the Hospital Frailty Risk Score (HFRS), and a four-item modified Frailty Index (mFI-4; the mFI-5 without functional status) described in Methods.

| Variable | Level | Overall | Female | Male | p-value | FDR |
| --- | --- | --- | --- | --- | --- | --- |
| **n** |  | 10719 | 3689 | 7030 |  |  |
| **Age median [Q1,Q3]** |  | 66.0 [55.0,74.0] | 66.0 [53.0,75.0] | 66.0 [56.0,73.0] | 0.467 | 0.467 |
| **Age group n (%)** | 0-17 | 367 (3.4) | 175 (4.7) | 192 (2.7) | <0.001 | 0.0016 |
|  | 18-44 | 941 (8.8) | 408 (11.1) | 533 (7.6) |  |  |
|  | 45-64 | 3715 (34.7) | 1179 (32.0) | 2536 (36.1) |  |  |
|  | 65-79 | 4684 (43.7) | 1470 (39.8) | 3214 (45.7) |  |  |
|  | 80+ | 1012 (9.4) | 457 (12.4) | 555 (7.9) |  |  |
| **ASA score n (%)** | 1 | 1227 (11.4) | 630 (17.1) | 597 (8.5) | <0.001 | 0.0016 |
|  | 2 | 213 (2.0) | 78 (2.1) | 135 (1.9) |  |  |
|  | 3 | 3803 (35.5) | 1549 (42.0) | 2254 (32.1) |  |  |
|  | 4 | 5476 (51.1) | 1432 (38.8) | 4044 (57.5) |  |  |
| **Physiological reserve n (%)** | Frail | 6139 (57.3) | 1666 (45.2) | 4473 (63.6) | <0.001 | 0.0016 |
|  | Pre-frail | 3558 (33.2) | 1501 (40.7) | 2057 (29.3) |  |  |
|  | Non-frail | 1022 (9.5) | 522 (14.2) | 500 (7.1) |  |  |
| **Emergency surgery n (%)** | No | 9250 (86.3) | 3221 (87.3) | 6029 (85.8) | 0.028 | 0.0373 |
|  | Yes | 1469 (13.7) | 468 (12.7) | 1001 (14.2) |  |  |
| **Surgery duration median [Q1,Q3]** |  | 213.0 [147.0,281.0] | 193.0 [135.0,267.0] | 221.0 [156.0,285.0] | <0.001 | 0.0016 |
| **Procedure cluster n (%)** | NervousSystem (c1) | 903 (8.4) | 365 (9.9) | 538 (7.7) | <0.001 | 0.0016 |
|  | Thoracic (c3) | 807 (7.5) | 317 (8.6) | 490 (7.0) |  |  |
|  | Cardiac (c4) | 3764 (35.1) | 836 (22.7) | 2928 (41.7) |  |  |
|  | Vascular (c5) | 769 (7.2) | 189 (5.1) | 580 (8.3) |  |  |
|  | AbdominalWall (c6) | 530 (4.9) | 249 (6.7) | 281 (4.0) |  |  |
|  | GastroIntestinal (c7) | 298 (2.8) | 105 (2.8) | 193 (2.7) |  |  |
|  | Hepatobiliary (c8) | 306 (2.9) | 95 (2.6) | 211 (3.0) |  |  |
|  | UroAndrological (c10) | 346 (3.2) | 98 (2.7) | 248 (3.5) |  |  |
|  | Gynecological (c11) | 303 (2.8) | 303 (8.2) | 0 (0.0) |  |  |
|  | Musculoskeletal (c12) | 1623 (15.1) | 711 (19.3) | 912 (13.0) |  |  |
|  | Cutaneous (c13) | 213 (2.0) | 95 (2.6) | 118 (1.7) |  |  |
|  | Unclassified | 857 (8.0) | 326 (8.8) | 531 (7.6) |  |  |
| **postoperative infection n (%)** | No | 10215 (95.3) | 3499 (94.8) | 6716 (95.5) | 0.123 | 0.141 |
|  | Yes | 504 (4.7) | 190 (5.2) | 314 (4.5) |  |  |

### Discrimination was similar by sex but heterogeneous by age and physiological reserve

Crude infection prevalence was similar between sexes and marginally higher in women (5.2% vs 4.5%). These findings indicate that recorded sex was correlated with age, physiological reserve, and procedure cluster. Age and physiological reserve were the main audited axes with deployment-relevant performance differences, while procedure cluster served as a case-mix reference axis.

Overall discrimination was strong and nearly identical by sex (Figure 1A, 1D): female-minus-male ΔAUROC was +0.004 (95% CI −0.029 to +0.037). At t = 0.10, sensitivity was 0.900 in women and 0.898 in men, and specificity was 0.640 versus 0.650. Sex-specific gaps were small: equal opportunity 0.002, predictive equality 0.010, equalized odds 0.010, and statistical parity 0.014 (Table 2). Discrimination varied more by age than by sex (Figure 1B, Table 2). AUROC was 0.807 at ages 0-17 and 0.805 at ≥80 years, versus 0.888 to 0.889 at ages 45-79. At t = 0.10, specificity was lowest at the age extremes (0.462 and 0.577), versus 0.654 to 0.682 at ages 18-79. The age equalized-odds gap was 0.220, mainly from specificity differences, so a single global threshold flagged more non-infected patients at the age extremes.

**Figure 1.**
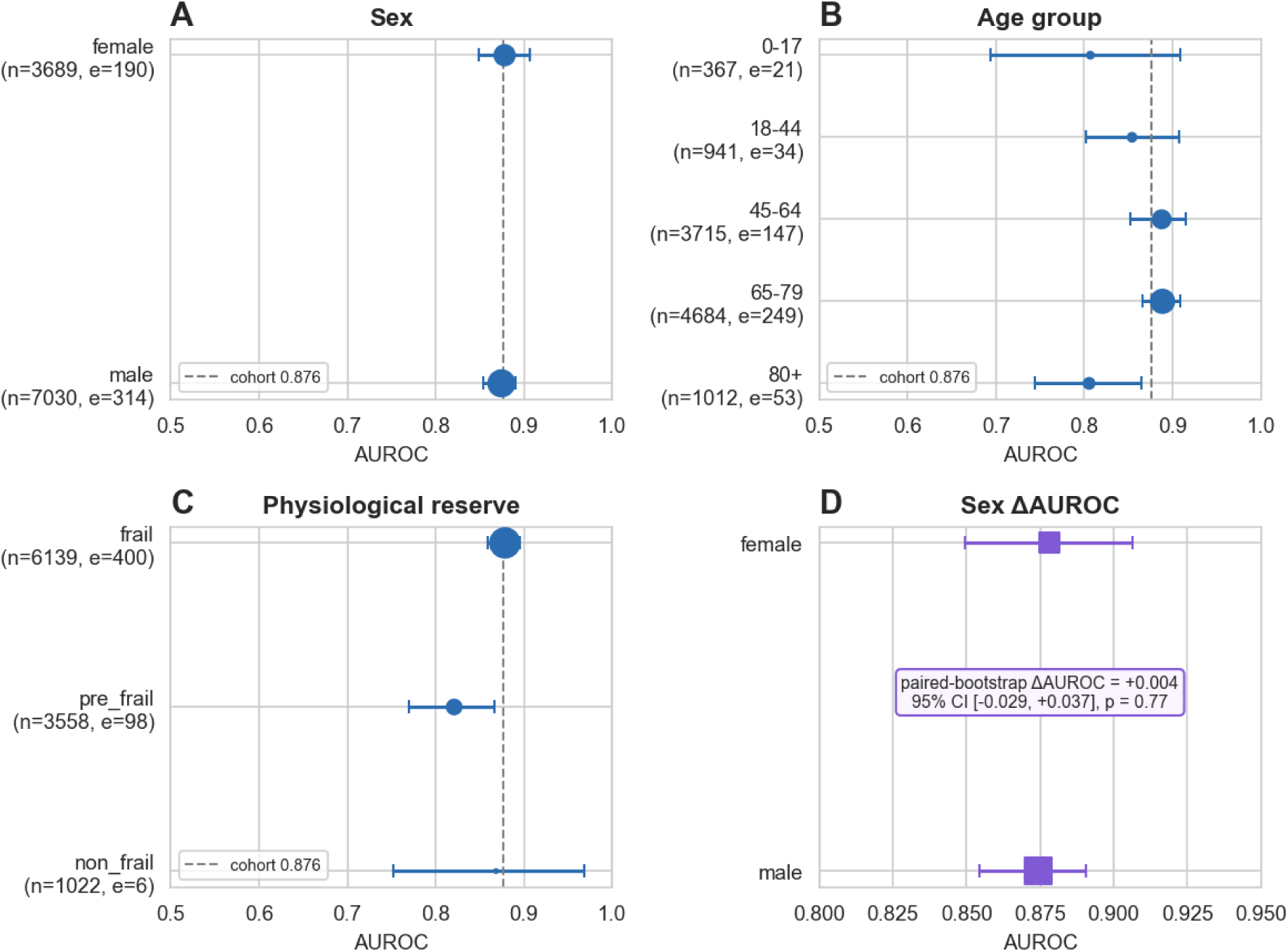
Subgroup discrimination. Panels A–C: forest plots of within-subgroup AUROC with 95% bootstrap confidence intervals (200 stratified resamples) across (A) sex (female, male), (B) age stratum (0–17, 18–44, 45–64, 65–79, ≥80 years), and (C) physiological-reserve composite (frail, pre-frail, non-frail). Point sizes are proportional to within-subgroup event count; the vertical dashed line marks the overall cohort AUROC of 0.876 (95% CI 0.860–0.890). (D) Female and male AUROC estimates on an expanded scale, with the paired-bootstrap ΔAUROC annotation for the sex contrast (female minus male: +0.004; 95% CI −0.029 to +0.037; p = 0.77, two-sided percentile test). The paired contrast is shown for sex only because it is the single binary axis, where one female-versus-male difference is well defined and the near-identical performance is a primary claim.

**Table 2.** Per-subgroup discrimination on the audited cohort (n = 10,719; 504 infection events). Within-subgroup AUROC, AUPRC, sensitivity, and specificity at the t = 0.10 operating threshold, with 95% bootstrap CIs (200 resamples). Gap statistics per axis are reported in the section header rows: EOD = equal-opportunity difference, the sensitivity gap; PE = predictive-equality difference, the false-positive-rate gap; EOG = equalized-odds gap, the larger of the sensitivity and false-positive-rate gaps; and SP = statistical-parity difference, the difference in the proportion of patients flagged.

| Subgroup | n | Events (%) | AUROC (95% CI) | AUPRC (95% CI) | Sensitivity (95% CI) | Specificity (95% CI) |
| --- | --- | --- | --- | --- | --- | --- |
| **Sex**<br>EOD 0.002 ·<br>PE 0.010 ·<br>EOG 0.010 ·<br>SP 0.014 |  |  |  |  |  |  |
| **Overall** | 10,719 | 504 (4.7) | 0.876 (0.860–0.890) | 0.376 (0.337–0.414) | 0.899 (0.871–0.923) | 0.647 (0.639–0.658) |
| **Female** | 3,689 | 190 (5.2) | 0.878 (0.850–0.906) | 0.408 (0.355–0.484) | 0.900 (0.853–0.942) | 0.640 (0.626–0.655) |
| **Male** | 7,030 | 314 (4.5) | 0.875 (0.854–0.891) | 0.356 (0.311–0.412) | 0.898 (0.863–0.930) | 0.650 (0.639–0.662) |
| **Age group**<br>EOD 0.095 ·<br>PE 0.220 ·<br>EOG 0.220 ·<br>SP 0.220 |  |  |  |  |  |  |
| **Overall** | 10,719 | 504 (4.7) | 0.876 (0.860–0.895) | 0.376 (0.336–0.425) | 0.899 (0.875–0.921) | 0.647 (0.638–0.657) |
| **0-17** | 367 | 21 (5.7) | 0.807 (0.694–0.909) | 0.246 (0.146–0.429) | 0.857 (0.667–1.000) | 0.462 (0.413–0.512) |
| **18-44** | 941 | 34 (3.6) | 0.854 (0.802–0.908) | 0.329 (0.200–0.482) | 0.824 (0.706–0.941) | 0.682 (0.654–0.710) |
| **45-64** | 3,715 | 147 (4.0) | 0.888 (0.853–0.915) | 0.382 (0.309–0.467) | 0.918 (0.871–0.959) | 0.666 (0.649–0.682) |
| **65-79** | 4,684 | 249 (5.3) | 0.889 (0.866–0.909) | 0.438 (0.368–0.506) | 0.912 (0.876–0.944) | 0.654 (0.641–0.669) |
| **80+** | 1,012 | 53 (5.2) | 0.805 (0.744–0.865) | 0.227 (0.153–0.334) | 0.849 (0.755–0.943) | 0.577 (0.545–0.608) |
| **Physiological reserve**<br>EOD 0.142 ·<br>PE 0.232 ·<br>EOG 0.232 ·<br>SP 0.262 |  |  |  |  |  |  |
| **Overall** | 10,719 | 504 (4.7) | 0.876 (0.858–0.893) | 0.376 (0.337–0.420) | 0.899 (0.869–0.925) | 0.647 (0.637–0.657) |
| **Frail** | 6,139 | 400 (6.5) | 0.878 (0.859–0.895) | 0.428 (0.384–0.473) | 0.927 (0.900–0.950) | 0.584 (0.572–0.596) |
| **Pre-frail** | 3,558 | 98 (2.8) | 0.821 (0.770–0.867) | 0.190 (0.143–0.275) | 0.786 (0.694–0.867) | 0.702 (0.689–0.717) |
| **Non-frail** | 1,022 | 6 (0.6) | 0.868 (0.752–0.969) | 0.084 (0.020–0.358) | 0.833 (0.500–1.000) | 0.816 (0.795–0.843) |

Discrimination was lowest in pre-frail patients (Figure 1C, Table 2); the non-frail estimate was imprecise because this stratum contained only six infection events. At t = 0.10, the share of true infections correctly flagged (sensitivity) was lowest in pre-frail patients, where the model missed 21% of infections, compared with 7% in frail patients. The corresponding sensitivity gap (equal-opportunity difference) was 0.142 and the equalized-odds gap was 0.232. The procedure-cluster reference axis showed an even larger raw discrimination gap (equalized-odds gap 0.516; Supplementary Table 5), but this gap mostly reflects that different operations carry different infection risks and that some clusters had very few events, rather than a fairness problem in the model.

### Calibration and clinical utility by subgroup

Calibration was the main subgroup failure mode (Figure 2, Table 3). The calibration curves showed that the model overestimated risk across audited subgroups, most strongly in patients with preserved physiological reserve (Figure 2A). Quantifying this, the frail-versus non-frail calibration-in-the-large gap was 1.62 log-odds units (95% CI 0.93 to 2.69), the largest gap on any axis (Figure 2B). Sex and procedure-cluster calibration gaps were small (Supplementary Table 6), indicating that physiological-reserve-related miscalibration was not explained by procedure cluster alone. This miscalibration changed the clinical utility of the score (Figure 2C, 2D): at t = 0.10, net benefit was positive overall and in frail patients (about +17 net infections identified per 1,000) but negative in pre-frail and non-frail patients (about −11 and −15 per 1,000) and in patients under 45. Re-expressed as natural frequencies (Figure 3), the model flagged 188 per 1,000 non-frail patients at t = 0.10 even though only about 5 per 1,000 developed infections. Across-threshold net-benefit gaps were largest for physiological reserve and procedure cluster and negligible for recorded sex.

**Figure 2.**
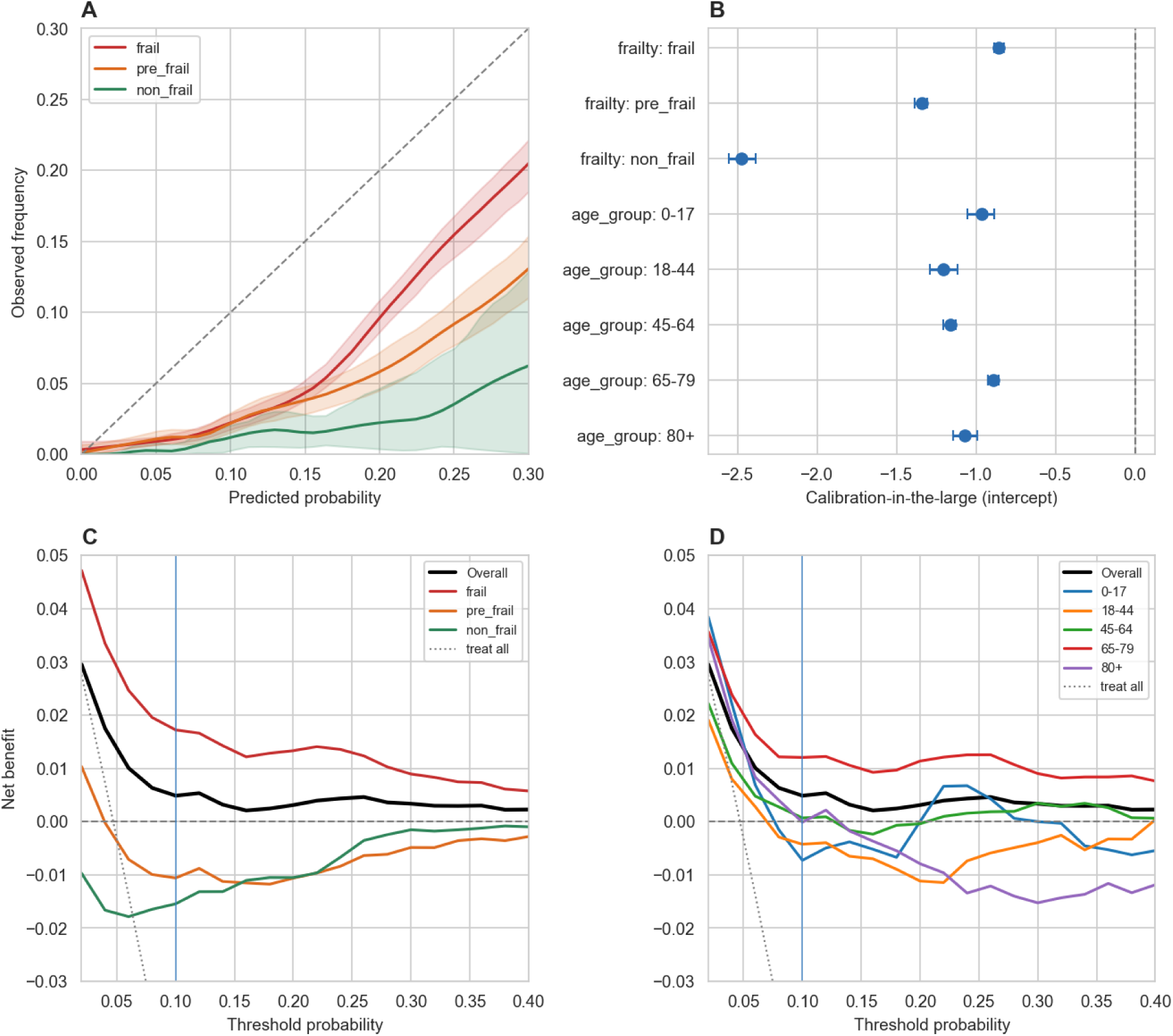
Subgroup calibration and clinical utility for the two axes that drive calibration heterogeneity (physiological reserve and age). (A) LOESS-smoothed calibration curves with 95% bootstrap bands (200 stratified resamples) by physiological reserve (frail, pre-frail, non-frail), over the predicted-probability range [0, 0.30]; the dashed diagonal is perfect calibration and curves below it indicate overestimation. (B) Calibration-in-the-large (fixed-slope logit intercept) with 95% bootstrap CIs for the physiological-reserve and age strata; 0 is perfect calibration and negative values indicate overestimation. (C, D) Decision curves of net benefit across t = 0.02 to 0.40 for (C) physiological reserve and (D) age. The t = 0.10 operating threshold (vertical blue line) is the risk level at which intensified monitoring would be triggered. "Treat-all" (dotted gray) is the policy of applying intensified monitoring to every patient regardless of predicted risk, equivalent to a threshold of 0; the zero line is "treat-none," meaning no intensified monitoring based on the score. Here, intensified monitoring denotes closer clinical surveillance, for example more frequent vital-sign and laboratory checks and earlier clinical review, rather than measurement with a specific device or treatment escalation. A useful model lies above both reference strategies.

**Figure 3.**
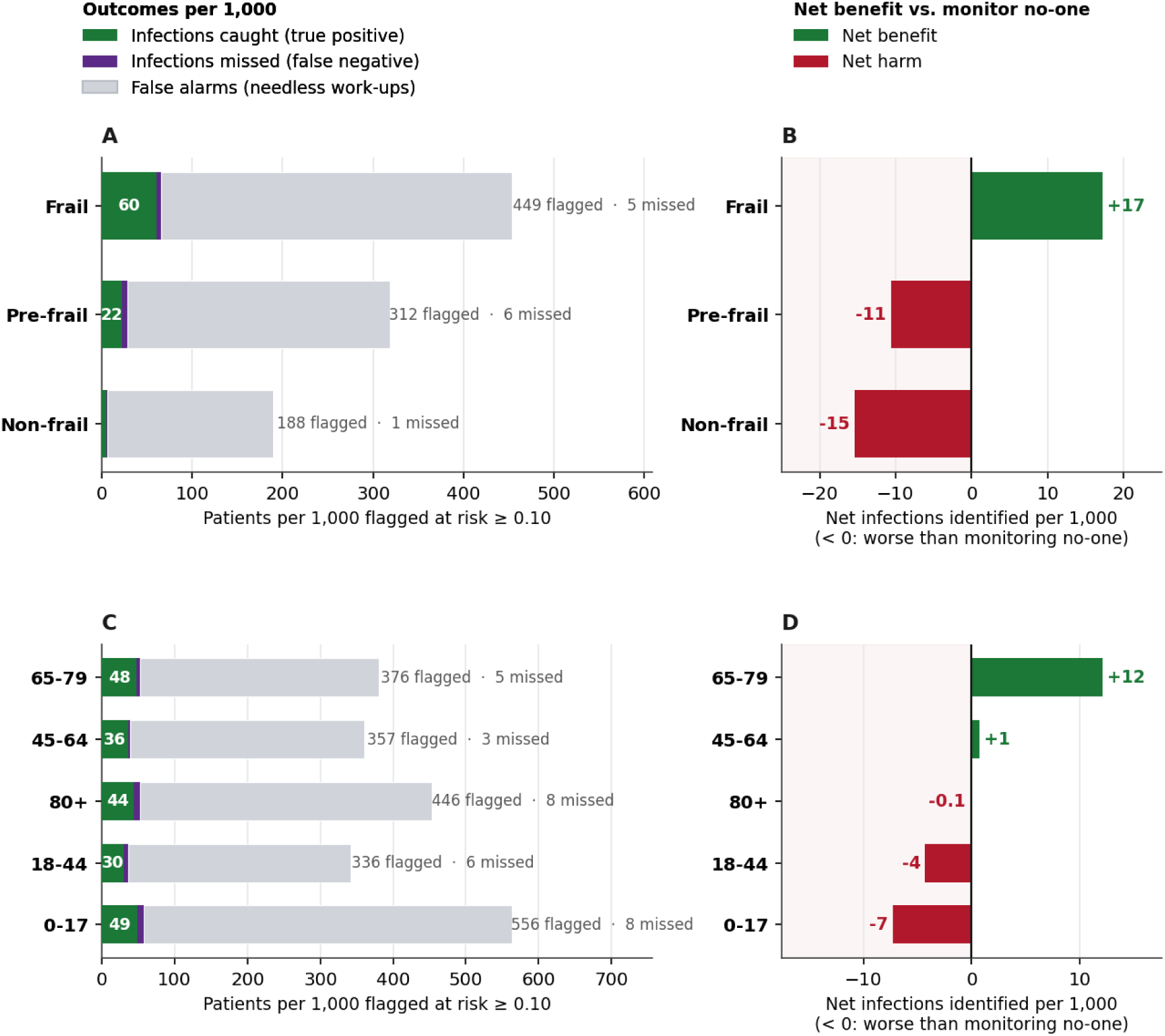
Clinical consequences of subgroup miscalibration at the 0.10 monitoring threshold, per 1,000 patients. Natural-frequency re-expression of audited operating characteristics and decision-curve net benefit. (A, C) Patients flagged at predicted risk ≥ 0.10 decomposed into infections caught (green), infections missed (purple), and false alarms triggering unnecessary intensified monitoring (grey), by physiological reserve (A) and age (C); labels give total flagged and missed. (B, D) Net benefit versus a monitor-no-one strategy, as net infections identified per 1,000, by reserve (B) and age (D); values below zero (red) indicate net harm. Subgroups ordered by net benefit.

**Table 3.** Per-subgroup calibration of the end-of-surgery infection prediction model for the three prespecified subgroup axes. Within-subgroup calibration intercept (calibration-in-the-large; fixed-slope logit recalibration) and calibration slope (logistic regression of outcome on linear predictor) with 95% bootstrap CIs (200 stratified resamples). Brier score with 95% CI, expected calibration error (ECE; 10 quantile bins), and integrated calibration index (ICI). Per-axis maximum intercept- and slope-gaps with 95% bootstrap CIs are reported in the section header rows.

| Subgroup | n | Events (%) | Intercept (95% CI) | Slope (95% CI) | Brier (95% CI) | ECE | ICI |
| --- | --- | --- | --- | --- | --- | --- | --- |
| **Sex** |  |  |  |  |  |  |  |
| **max ΔIntercept 0.09 (0.01, 0.28)** |  |  |  |  |  |  |  |
| **max ΔSlope 0.01 (0.00, 0.26)** |  |  |  |  |  |  |  |
| **Overall** | 10,719 | 504 (4.7) | -1.02 (-1.04, -0.99) | 1.57 (1.46, 1.68) | 0.041 (0.040, 0.042) | 0.062 | 0.061 |
| **Female** | 3,689 | 190 (5.2) | -0.96 (-1.01, -0.92) | 1.56 (1.38, 1.80) | 0.043 (0.041, 0.045) | 0.061 | 0.06 |
| **Male** | 7,030 | 314 (4.5) | -1.05 (-1.09, -1.02) | 1.57 (1.43, 1.71) | 0.040 (0.039, 0.041) | 0.063 | 0.061 |
| **Age group** |  |  |  |  |  |  |  |
| **max ΔIntercept 0.32 (0.23, 0.83)** |  |  |  |  |  |  |  |
| **max ΔSlope 0.51 (0.29, 1.32)** |  |  |  |  |  |  |  |
| **Overall** | 10,719 | 504 (4.7) | -1.02 (-1.05, -1.00) | 1.57 (1.47, 1.71) | 0.041 (0.040, 0.042) | 0.062 | 0.061 |
| **0-17** | 367 | 21 (5.7) | -0.96 (-1.05, -0.89) | 1.54 (0.92, 2.45) | 0.055 (0.050, 0.060) | 0.073 | 0.071 |
| **18-44** | 941 | 34 (3.6) | -1.21 (-1.29, -1.11) | 1.40 (1.05, 1.84) | 0.036 (0.032, 0.040) | 0.065 | 0.064 |
| **45-64** | 3,715 | 147 (4.0) | -1.16 (-1.21, -1.12) | 1.59 (1.39, 1.86) | 0.036 (0.034, 0.038) | 0.065 | 0.063 |
| **65-79** | 4,684 | 249 (5.3) | -0.89 (-0.92, -0.86) | 1.68 (1.50, 1.88) | 0.042 (0.041, 0.044) | 0.057 | 0.056 |
| **80+** | 1,012 | 53 (5.2) | -1.07 (-1.14, -0.99) | 1.16 (0.88, 1.51) | 0.053 (0.049, 0.057) | 0.074 | 0.075 |
| **Physiological reserve** |  |  |  |  |  |  |  |
| **max ΔIntercept 1.62 (0.93, 2.69)** |  |  |  |  |  |  |  |
| **max ΔSlope 0.31 (0.16, 1.71)** |  |  |  |  |  |  |  |
| **Overall** | 10,719 | 504 (4.7) | -1.02 (-1.04, -0.99) | 1.57 (1.46, 1.71) | 0.041 (0.040, 0.042) | 0.062 | 0.061 |
| **Frail** | 6,139 | 400 (6.5) | -0.86 (-0.88, -0.83) | 1.59 (1.47, 1.73) | 0.052 (0.050, 0.054) | 0.064 | 0.062 |
| **Pre-frail** | 3,558 | 98 (2.8) | -1.34 (-1.38, -1.31) | 1.28 (1.09, 1.54) | 0.031 (0.029, 0.032) | 0.062 | 0.062 |
| **Non-frail** | 1,022 | 6 (0.6) | -2.47 (-2.56, -2.39) | 1.55 (0.79, 2.91) | 0.011 (0.010, 0.013) | 0.055 | 0.055 |

### Post hoc mitigation: testing isotonic recalibration versus score-distribution postprocessing

Group-specific isotonic recalibration and Wasserstein-barycenter score postprocessing were evaluated as post hoc score-level mitigation strategies (Figure 4, Table 4, Supplementary Table 9). Group-specific isotonic recalibration was most consistent for recorded sex and physiological reserve (Figure 4A and 4B): calibration-in-the-large moved to approximately zero in all reserve categories and both sex strata, and Brier score improved in each (Table 4). This reduced average physiological-reserve-stratified overestimation, although some estimates remained statistically unstable: the non-frail group contained only six events, and its recalibrated slope fell to 0.07 despite correction of average risk.

**Figure 4.**
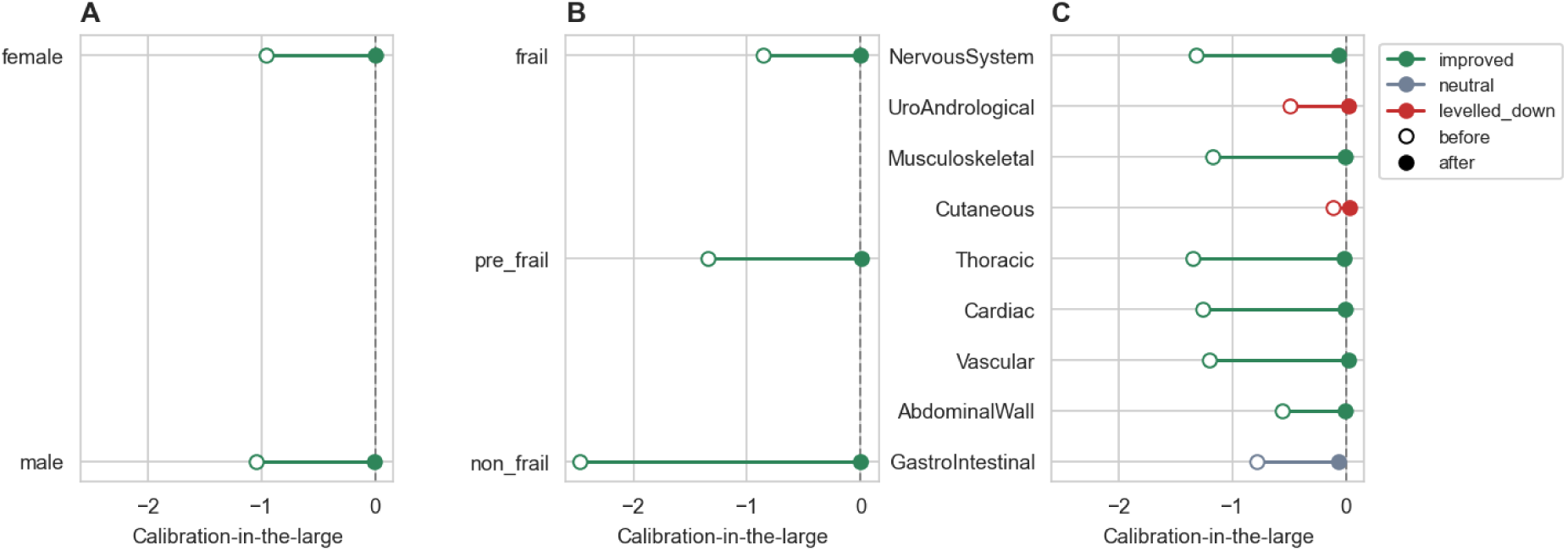
Calibration-in-the-large per subgroup, before and after isotonic recalibration. (A) Sex. (B) Physiological reserve. (C) Procedure cluster (reference axis). Open circles = before recalibration; filled circles = after. Markers are colored by the audit’s leveling-down flag (green = improved, gray = neutral, red = leveled down). The dashed vertical line at 0 marks perfect calibration-in-the-large. Values close to 0 indicate correction of average calibration; calibration slope and Brier score in Table 4 should be used to assess stability and possible overfitting.

**Table 4.** Within-group isotonic recalibration of the end-of-surgery infection prediction model, before vs after, by sensitive axis. Group-specific isotonic recalibration was fitted independently within each subgroup of the sex, physiological-reserve, and procedure-cluster axes. For each subgroup the Brier score, calibration-in-the-large (intercept; 0 = perfectly calibrated), and calibration slope (1 = perfectly calibrated) are reported before and after recalibration. The Flag column is the audit’s leveling-down classifier, which flags a subgroup as leveled down when the post-mitigation Brier score does not improve and calibration-in-the-large overshoots past zero.

| Subgroup | n | Events | Brier | Intercept | Slope | Flag |
| --- | --- | --- | --- | --- | --- | --- |
| **Sex** |  |  |  |  |  |  |
| **Female** | 3,689 | 190 | 0.043 → 0.039 | -0.96 → 0.00 | 1.56 → 0.86 | improved |
| **Male** | 7,030 | 314 | 0.040 → 0.035 | -1.05 → 0.00 | 1.57 → 0.88 | improved |
| **Physiological reserve** |  |  |  |  |  |  |
| **Frail** | 6,139 | 400 | 0.052 → 0.047 | -0.86 → 0.00 | 1.59 → 0.89 | improved |
| **Pre-frail** | 3,558 | 98 | 0.031 → 0.025 | -1.34 → 0.01 | 1.28 → 0.81 | improved |
| **Non-frail** | 1,022 | 6 | 0.011 → 0.006 | -2.47 → 0.00 | 1.55 → 0.07 | improved |
| **Procedure cluster** |  |  |  |  |  |  |
| **NervousSystem (c1)** | 903 | 22 | 0.023 → 0.021 | -1.32 → -0.06 | 1.83 → 0.14 | improved |
| **Thoracic (c3)** | 807 | 22 | 0.026 → 0.021 | -1.35 → -0.02 | 1.94 → 0.19 | improved |
| **Cardiac (c4)** | 3,764 | 118 | 0.032 → 0.026 | -1.27 → 0.00 | 1.58 → 0.85 | improved |
| **Vascular (c5)** | 769 | 30 | 0.036 → 0.031 | -1.20 → 0.02 | 1.73 → 0.50 | improved |
| **AbdominalWall (c6)** | 530 | 67 | 0.078 → 0.076 | -0.57 → 0.00 | 1.62 → 0.46 | improved |
| **GastroIntestinal (c7)** | 298 | 36 | 0.090 → 0.089 | -0.79 → -0.06 | 1.75 → 0.16 | neutral |
| **UroAndrological (c10)** | 346 | 28 | 0.064 → 0.067 | -0.49 → 0.03 | 1.36 → 0.11 | leveled down |
| **Musculoskeletal (c12)** | 1,623 | 62 | 0.040 → 0.035 | -1.17 → 0.00 | 1.34 → 0.37 | improved |
| **Cutaneous (c13)** | 213 | 43 | 0.127 → 0.134 | -0.11 → 0.04 | 1.38 → 0.24 | leveled down |

Wasserstein-barycenter postprocessing reduced the targeted false-positive-rate gap across procedure clusters but worsened calibration in most audited clusters (Supplementary Table 9). This contrast is expected because Wasserstein postprocessing equalizes score distributions rather than observed-versus-predicted risk. Thus, enforcing score-distribution parity reduced selected parity gaps but worsened calibration, making it less appropriate than isotonic recalibration for the observed failure mode.

### Intersectional and secondary interaction analyses

The secondary analyses were prespecified model-based interaction tests (sex by physiological reserve, sex by ASA, and a sex by comorbidity screen) examining whether covariate-outcome associations differed by sex. Pairwise intersectional analyses were limited by sparse cells (Supplementary Table 7), but the patterns mirrored the age and physiological-reserve findings. Equalized-odds gaps were largest for age by physiological reserve. Intersectional analyses did not identify an additional pattern beyond the prespecified age and physiological-reserve margins.

The sex-by-physiological-reserve interaction term for infection was statistically significant but small (likelihood-ratio p = 0.018, ΔR² = 0.0008). As Table 1 shows, sex was associated with physiological reserve and case mix, but infection prevalence itself was similar between sexes, so the interaction reflects composition rather than a sex difference in infection risk.

Secondary sex-stratified analyses (Figure 5, Supplementary Table 8) supported interpretation of recorded sex as a compositional rather than a primary performance axis. Men and women differed substantially in procedure mix and baseline acuity (Figure 5A). However, comorbidity-specific infection associations did not show a consistent sex-specific direction (Figure 5B), and within-cluster AUROC differences varied in both directions rather than indicating systematic degradation for either sex (Figure 5C). Infection risk increased with ASA physical status in both sexes, with a statistically significant but small sex-by-ASA interaction (Figure 5D). These findings indicate that recorded sex captured differences in case mix and baseline clinical composition, whereas the deployment-relevant performance failures were concentrated along age and physiological-reserve axes rather than sex-marginal model performance.

**Figure 5.**
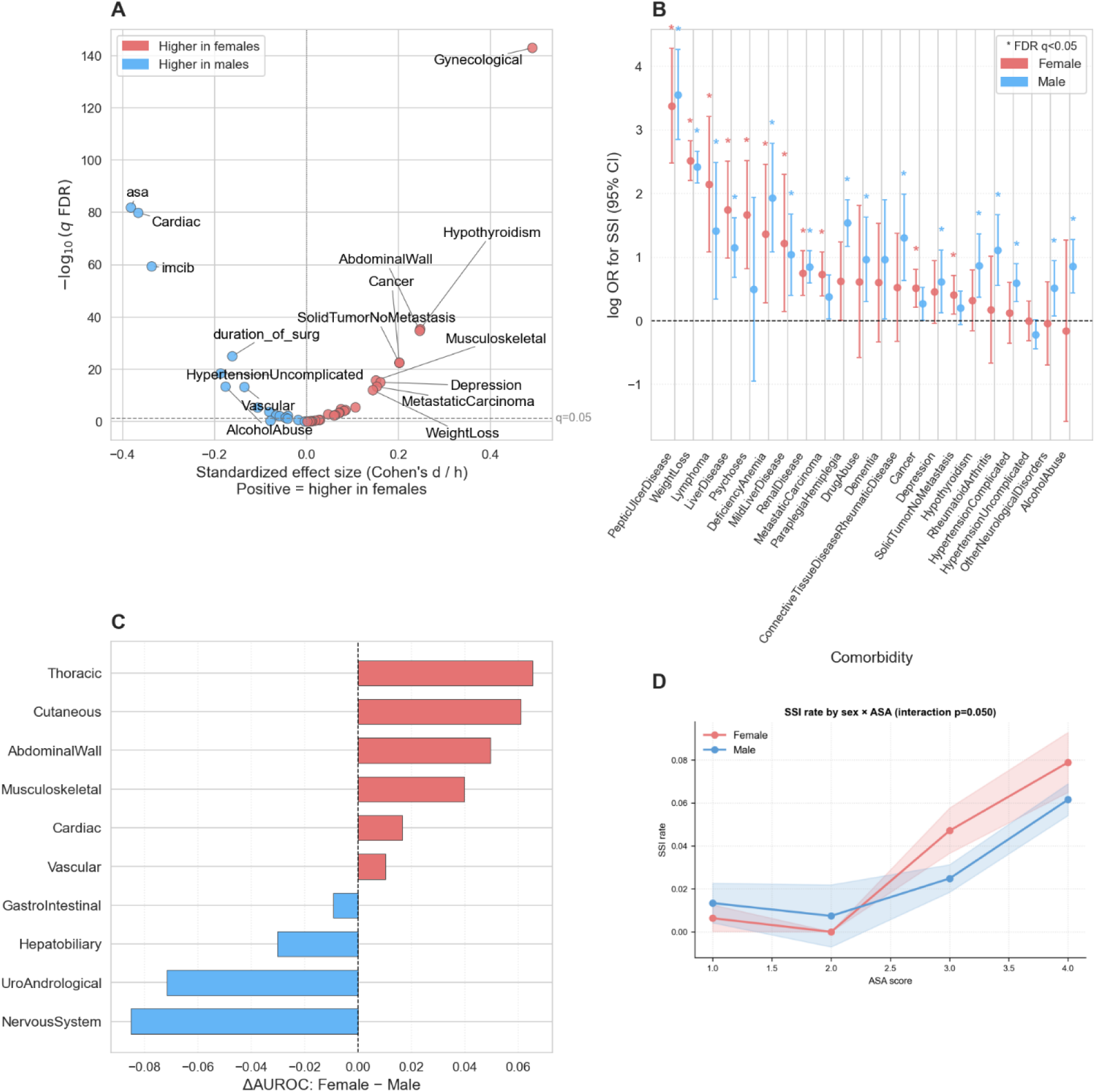
Targeted characterization of the sex axis. (A) Feature-by-sex effect-size volcano: standardized effect size (Cohen’s d for continuous features, Cohen’s h for binary features) against −log₁₀(FDR-corrected q) for every candidate feature; positive effect = higher value or prevalence in females, points colored by direction, the dashed line marks q = 0.05, and features with |effect| ≥ 0.15 or strong significance are labelled. (B) Per-comorbidity odds ratio for postoperative bacterial infection by sex on the log scale with 95% CI, estimated separately within females (salmon) and males (blue); comorbidities are ordered by the female OR and * marks FDR q < 0.05. (C) Per-cluster AUROC gap: difference in model discrimination (female − male AUROC) within each surgical procedure cluster from out-of-fold predictions, positive = better discrimination in females. (D) Sex × ASA interaction: observed postoperative infection rate by ASA physical-status score stratified by sex, with 95% CI bands and the likelihood-ratio interaction p-value.

### Portability analysis

In the prespecified SUPPORT2 portability analysis, the *isitfair* workflow was applied to a logistic-regression model for in-hospital mortality in 1,821 held-out critically ill US adults (mortality 25.9%) and reproduced the same subgroup-reporting sequence used in our cohort: subgroup discrimination, calibration, threshold-specific performance, and clinical-utility summaries. It also exercised a race/ethnicity axis that was unavailable in the Swiss EHR.

This analysis was a workflow demonstration rather than external validation of the perioperative infection model, and SUPPORT2 differs from our cohort in population, outcome, and prevalence, so results are not directly comparable across the two datasets. Within SUPPORT2, recorded-sex AUROC differed by about 0.02 (0.833 in females vs. 0.856 in males), whereas AUROC varied from 0.811 to 0.879 across age strata and from 0.800 to 0.855 across race/ethnicity groups, although the extreme estimates were based on <60 patients. The same calibration- and utility-reporting steps ran end to end; the full generated report is provided in the Supplementary Materials.

## DISCUSSION

This audit identified a subgroup-specific deployment risk that would have been missed by discrimination-only or sex-marginal evaluation. The main failure was not poor average ranking, but physiological-reserve-stratified miscalibration that changed the clinical utility of the end-of-surgery infection-risk score. This pattern supports subgroup fairness auditing as a deployment-readiness assessment of discrimination, calibration, clinical utility, and mitigation effects, rather than a checklist of demographic error-rate gaps alone.

### Discrimination, calibration, and clinical utility

Discrimination heterogeneity was clinically relevant but less dominant than calibration. AUROC fell at both age extremes and in pre-frail patients (Figure 1, Table 2). Because event prevalence differed across strata, AUROC alone can be misleading; per-subgroup results should be interpreted alongside prevalence, not ranking metrics alone [46]. A single operating threshold can behave differently when ranking performance and prevalence differ, and sparse-stratum threshold estimates are unstable. A safer deployment strategy is to recalibrate probabilities within clinically justified strata, then prospectively define subgroup-specific criteria before use [21,22,44,47].

Calibration was critical because the end-of-surgery infection-risk score would be used as an individualized probability at a predefined monitoring threshold. The physiological-reserve-stratified failure was mainly calibration-in-the-large, showing why subgroup intercepts and slopes should accompany binned and curve-based summaries [21,34–37]. Decision-curve analysis then translated this miscalibration into deployment risk: systematic overprediction can make model-guided monitoring worse than monitoring no one, especially in patients with high physiological reserve (the non-frail stratum), where unnecessary monitoring escalation is less likely to be offset by clinical gain [38,48–50].

### Mitigation, leveling down, and practical deployment

Mitigation was evaluated under a prespecified leveling-down criterion, defined here as an apparent parity improvement that reduces subgroup gaps by worsening absolute performance in one or more subgroups. Smaller parity gaps are not desirable if they arise from degraded subgroup performance. Isotonic recalibration and Wasserstein-barycenter postprocessing are monotonic within subgroups and therefore largely preserve rank ordering [41], but optimize different objectives. Isotonic recalibration corrects predicted-versus-observed mismatch, whereas Wasserstein postprocessing equalizes score distributions and can reduce a parity gap while worsening calibration (Table 4, Supplementary Table 9). Thus, isotonic recalibration matched the observed calibration-driven failure. For deployment, it should be considered only when a subgroup has clear calibration error (intercept or slope materially departing from 0 or 1 with CIs excluding the target) and enough calibration-set events to fit the transform. In sparse strata, average calibration correction can coexist with unstable slopes, so parametric or hierarchical recalibration may be safer [19,40,51–53]. Neither postprocessing approach can repair true discrimination loss. The weaker separation in the youngest, oldest, and pre-frail patients requires threshold review and larger validation cohorts [21,34]. The original model used SMOTE within training folds, which can shift predicted probabilities in low-prevalence strata [54]. Because a refit without SMOTE was not available for this audit, the contribution of class rebalancing to subgroup miscalibration remains unresolved. The deployment implication is unchanged: probabilities require direct subgroup evaluation rather than inference from discrimination alone.

### Intersectional and sex-composition findings

Pairwise intersectional analyses added little beyond the single-axis findings: the gaps mainly tracked the age and physiological-reserve margins, and the sex by physiological-reserve result reinforced the same point. Recorded sex was associated with physiological reserve and case mix, but sex-marginal performance was similar. An apparently reassuring sex-only audit can therefore conceal composition-mediated harm when sex correlates with the axis on which the model fails [9,10]. In this audit, physiological-reserve recalibration was the appropriate mitigation because it targeted the affected strata directly rather than using sex as a proxy axis [23,55].

### Related work

These findings extend previous clinical AI fairness work showing that aggregate calibration can coexist with subgroup deterioration [9] and that postoperative prediction models require explicit fairness evaluation [56]. They also support a practical distinction between parity-focused and calibration-focused mitigation: threshold-based parity criteria remain useful for describing error-rate gaps [32,33], but safe deployment requires predicted probabilities that match observed infection rates within each subgroup, not only balanced yes/no decisions.

### Equity and representation

Subgroup fairness auditing for deployment is also ethically motivated. Real-world EHR data can reveal deployment risks in the patient populations seen in practice, but equitable prediction requires systematic capture of protected and socially defined attributes. The absence of these variables limits the present audit, although the audit framework can accommodate these variables when they are available.

### Limitations

Several limitations should guide interpretation. The primary cohort came from a single Swiss tertiary center, and SUPPORT2 was a portability analysis of the audit workflow, not external validation of the infection model. Tertiary referral likely enriched complex, higher-acuity cases, increasing the frail stratum and shrinking preserved-reserve strata; the small non-frail event count therefore tempers interpretation of the largest calibration gap. Sex was administratively recorded as binary and did not capture gender identity. Race, ethnicity, migrant status, language, socioeconomic disadvantage, and rurality were unavailable. Systematic capture of socially defined attributes is required for equitable prediction. The physiological-reserve construct is an EHR-derived proxy, not a validated frailty instrument, and depends partly on diagnosis coding; the mFI-4 hypertension item used ICD-10 codes without medication linkage, potentially overcounting hypertension. Post-discharge infections may be missed by the index-admission ICD-10-GM outcome. Scores were out-of-fold cross-validation predictions rather than outputs from a locked deployment artifact, and mitigation was limited to post hoc score transformations rather than model retraining or prospective workflow redesign.

## CONCLUSION

Strong average discrimination and reassuring sex-marginal metrics are insufficient evidence for equitable clinical deployment. In this audit, only the joint evaluation of discrimination, calibration, and decision-curve utility exposed the deployment-relevant failure: physiological-reserve-stratified overprediction that reversed net benefit at the monitoring threshold. Before clinical use, the model would require subgroup-aware recalibration, prospective validation of the operating threshold, and prespecified subgroup-specific criteria for when the recalibrated score is considered fit for clinical use. More broadly, the *isitfair* workflow provides a transparent deployment-readiness framework for identifying subgroup failure, comparing mitigation strategies, and producing TRIPOD+AI-aligned reports for clinical prediction models.

## Data Availability

The Bern cohort contains patient-level electronic health record data and cannot be shared publicly. Aggregated subgroup outputs, analysis code, package versions, and the SUPPORT2 reproducibility example are available at https://github.com/HugoGuillen/isitfair.

## BACK MATTER

## Acknowledgments

We thank the members of the INFRA project, in particular Stefanie Marti. We thank Guy Maddern and Markus Trochsler for their valuable input.

## Funding

This research was supported by Swiss Personalized Health Network (SPHN) grant DEM-2022-08, the Multidisciplinary Center for Infectious Diseases (MCID) MA_18 of the University of Bern, and the Commission for Digitalization of the University of Bern.

## Competing interests

The authors declare no competing interests.

## Code availability

The *isitfair* package and analysis scripts are available at https://github.com/HugoGuillen/isitfair.

## Ethics approval

The study was approved by the Cantonal Ethics Committee of Bern (KEK-ID: 2021-00965), with a waiver of informed consent due to the study’s retrospective design.

## Author contributions

Hugo Guillen-Ramirez (Conceptualization, Data curation, Formal analysis, Investigation, Methodology, Software, Validation, Visualization, Writing—original draft, Writing—review & editing), Katharina Lucia Lucas (Investigation, Validation, Writing—review & editing), Yves Max Wintsch (Investigation, Validation, Writing—review & editing), Tobias Ueli Blatter (Data curation, Methodology, Writing—review & editing), Karen Triep (Conceptualization, Data curation, Writing—review & editing), Olga Endrich (Conceptualization, Data curation, Funding acquisition, Writing—review & editing), and Guido Beldi (Conceptualization, Funding acquisition, Investigation, Methodology, Project administration, Resources, Supervision, Validation, Writing—review & editing).

## Supplementary captions

**Supplementary Table 1. Sex-stratified effect-size scan of the cohort characterization variables (audited Bern cohort, *n* = 10,719; 504 infection events).** For each of the eight variables tested in Table 1, the table reports the female-vs-male effect size (Cohen’s d for continuous variables, Cramér’s V for categorical variables, or Cohen’s h for binary variables), its conventional magnitude class, the test p-value (Mann–Whitney U for continuous variables and χ² for categorical or binary variables), and the Benjamini–Hochberg FDR-adjusted q-value across the eight-variable family. Variables are ordered by ascending q-value.

**Supplementary Table 2. Code list and weights for the Hospital Frailty Risk Score (HFRS).** HFRS codes and weights follow Gilbert et al. (Lancet 2018; 391:1775–82), Supplementary Table A2 (109 categories, version 5.6). The admission score is the sum of the weights of all distinct categories present. Risk bands: low (score < 5; contributes "non-frail"), intermediate (5 ≤ score ≤ 15; contributes "pre-frail"), high (score > 15; contributes "frail").

**Supplementary Table 3. Item-to-code map for the modified Frailty Index (mFI-4).** The five-factor modified Frailty Index (mFI-5; Subramaniam et al., JAMA Surg 2018) includes functional status, which is not coded in our EHR. We therefore used the remaining four items (mFI-4). Each item scores 1 if any listed ICD-10 category is present; the item score is the sum (0–4).

**Supplementary Table 4. Composite physiological-reserve classification rule.** A patient is assigned the highest reserve-loss level any single instrument supports. Missing inputs are conservatively ignored (a criterion can only raise the reserve-loss level if its input is present).

**Supplementary Table 5. Per-subgroup discrimination on the audited cohort (*n* = 10,719; 504 infection events) for the case-mix reference axes (surgical specialty, procedure cluster) and the three prespecified pairwise intersections.** Within-subgroup AUROC, AUPRC, sensitivity, and specificity at the *t* = 0.10 operating threshold, with 95% bootstrap CIs (200 resamples). Gap statistics per axis are reported in the section header rows (EOD = equal-opportunity difference, PE = predictive-equality difference, EOG = equalized-odds gap, SP = statistical parity). Subgroups flagged with † used empirical-Bayes shrinkage toward the marginal rate (cell *n* events < 50).

**Supplementary Table 6. Per-subgroup calibration on the audited cohort (*n* = 10,719; 504 infection events) for the case-mix reference axes (surgical specialty, procedure cluster) and the three prespecified pairwise intersections.** Within-subgroup calibration intercept (calibration-in-the-large; fixed-slope logit recalibration) and calibration slope (logistic regression of outcome on linear predictor) with 95% bootstrap CIs (200 stratified resamples). Brier score and 95% CI expected calibration error (ECE; 10 quantile bins), and integrated calibration index (ICI). Per-axis maximum intercept- and slope-gaps with 95% bootstrap CIs are reported in the section header rows. Subgroups flagged with † used empirical-Bayes shrinkage toward the marginal rate (cell *n* events < 50).

**Supplementary Table 7. Two-way intersectional cell sparsity and gap statistics (audited Bern cohort, *n* = 10,719; 504 infection events).** For each of the three prespecified two-way intersections, every cell’s size, outcome count, and empirical-Bayes (EB) shrinkage status is reported. Cells with fewer than 50 outcome events (marked †) were estimated with EB shrinkage of the cell-level postoperative infection rate toward the marginal subgroup rate; the shrinkage factor (0 = no shrinkage, 1 = full shrinkage to the margin) is shown for those cells. The per-intersection header row gives the number of cells, the number shrunk, the equalized-odds gap (discrimination), and the maximum net-benefit gap with 95% bootstrap CI (clinical utility).

**Supplementary Table 8. Sex × comorbidity interaction screen for the postoperative infection outcome (audited Bern cohort, *n* = 10,719; 504 infection events).** For each of the 23 screened comorbidities, the odds ratio for postoperative infection (95% Wald CI) and the absolute postoperative infection rate among exposed vs unexposed patients are reported separately within each sex (the data plotted in Figure 5B), followed by the sex × comorbidity interaction: a likelihood-ratio test of nested logistic-regression models (adjusted for sex, age, ASA, and the comorbidity main effect) of whether the comorbidity → postoperative infection association differs between sexes, its Cox–Snell pseudo-*R*² increment (Δ*R*²), and the Benjamini–Hochberg FDR-adjusted *q*-value across the comorbidity family. Each interaction is a 1-degree-of-freedom likelihood-ratio test (binary comorbidity × binary sex). Rows are ordered by interaction *q*.

**Supplementary Table 9. Wasserstein-barycenter score postprocessing of the end-of-surgery infection prediction model on the procedure-cluster axis, before vs after (audited Bern cohort, *n* = 10,719; 504 infection events).** Wasserstein postprocessing was fitted independently across the procedure-cluster (surgical group) axis under a demographic-parity constraint: each cluster’s predicted-risk distribution is mapped, by a within-cluster quantile-to-quantile transformation, onto the group-size-weighted 1-D Wasserstein barycenter of the per-cluster calibration-set score distributions. The transform is monotone within each cluster, so within-cluster rank order (and AUROC) is preserved exactly; it equalizes the score distribution across clusters but does not target calibration. For each cluster the Brier score, calibration-in-the-large (intercept; 0 = perfectly calibrated), and calibration slope (1 = perfectly calibrated) are reported before and after postprocessing. The audit’s leveling-down classifier flags a cluster as *leveled down* when the post-mitigation Brier score does not improve. Low-event clusters 8 (Hepatobiliary) and 11 (Gynecological) and the unclassified bucket (together 1,466 patients, 76 events) did not meet the minimum per-group sample for a within-group fit; those patients retain their original predictions and are not shown.

**Supplementary Table 10. Clinical consequences of subgroup miscalibration at the 0.10 monitoring threshold, expressed as natural frequencies per 1,000 patients (audited Bern cohort, n = 10,719; 504 infection events).** For each subgroup of the three prespecified sensitive axes (physiological reserve, age stratum, recorded sex), the table reports, per 1,000 patients, the number flagged for intensified monitoring at the prespecified operating threshold t = 0.10 (predicted risk ≥ 0.10), decomposed into infections genuinely caught (true positives), false alarms (false positives), and infections missed (false negatives). These counts are derived directly from the audited per-subgroup outcome prevalence, sensitivity, and specificity at t = 0.10 (Table 2). Net benefit per 1,000 is the threshold-weighted count of net infections identified (true positives minus false positives weighted by the 1:9 harm trade-off implied by t = 0.10) and equals 1,000 times the decision-curve net benefit in Figure 2C–D; the standardized net benefit rescales this so that 1 is a hypothetical perfect model and values ≤ 0 indicate no benefit over monitoring no one. The verdict column flags a subgroup as net harm when its net benefit is negative, i.e. the score performs worse than monitoring no patient at this threshold.

## Supplementary methods

These Supplementary Methods give formal definitions, estimators, and equations underlying the fairness audit. They are the technical companion to the main-text Methods, which carries the design rationale and a plain-language description of each measure.

### Notation

The audited cohort has *n* patients indexed by *i* = 1, … , *n*. For each patient:

- *Y*_*i*_ ∈ {0,1} is the binary outcome (1 = bacterial postoperative infection within the index admission);
- *p*_*i*_ ∈ [0,1] is the model’s predicted probability that *Y*_*i*_ = 1, obtained as an out-of-fold prediction from 5-fold stratified cross-validation of the end-of-surgery model [6];
- *A*_*i*_ ∈ *G* is the value of the sensitive attribute (e.g. {female, male} for sex; {frail, pre-frail, non-frail} for physiological reserve).

For a subgroup *g*, let *I*_*g*_ = {*i*: *A*_*i*_ = *g*} be its patients, *n*_*g*_ =∣ *I*_*g*_ ∣ its size, 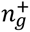 its event count, 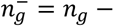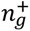 its non-event count, and 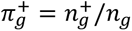 its outcome prevalence. Population-level quantities drop the *g* subscript. At a decision threshold *t* ∈ [0,1] a patient is flagged when *p*_*i*_ ≥ *t*. Within subgroup *g* the four standard counts are true positives TP_*g*_(*t*) (flagged events: *p*_*i*_ ≥ *t*, *Y*_*i*_ = 1), false positives FP_*g*_(*t*) (flagged non-events: *p*_*i*_ ≥ *t*, *Y*_*i*_ = 0), false negatives FN_*g*_(*t*) (missed events: *p*_*i*_ < *t*, *Y*_*i*_ = 1), and true negatives TN_*g*_(*t*) (correctly unflagged: *p*_*i*_ < *t*, *Y*_*i*_ = 0).

### Discrimination metrics

#### AUROC

The area under the receiver-operating-characteristic curve is the probability that the model assigns higher risk to a randomly chosen event case than to a randomly chosen non-event case in the same subgroup,

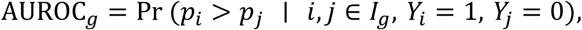

estimated by the normalized Mann–Whitney *U* statistic restricted to *I*_*g*_, with ties broken by averaging.

#### AUPRC

The area under the within-subgroup precision–recall curve, estimated as the trapezoidal sum over the operating points (TPR_*g*_(*t*), PPV_*g*_(*t*)) swept across all observed thresholds. It is the natural complement to AUROC at low prevalence.

#### Per-threshold rates

At the operating threshold *t* = 0.10 for intensifying postoperative-infection surveillance,

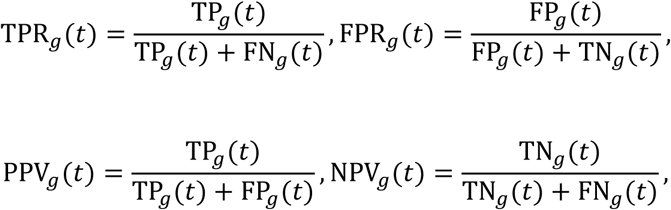

with specificity TNR_*g*_(*t*) = 1 − FPR_*g*_(*t*). Sensitivity (TPR) and specificity (TNR) are reported in the main discrimination tables; PPV and NPV are provided in the per-subgroup supplementary data files.

#### Gap statistics

For a pair of subgroups *g*, *g*^′^, all evaluated at the same *t*:

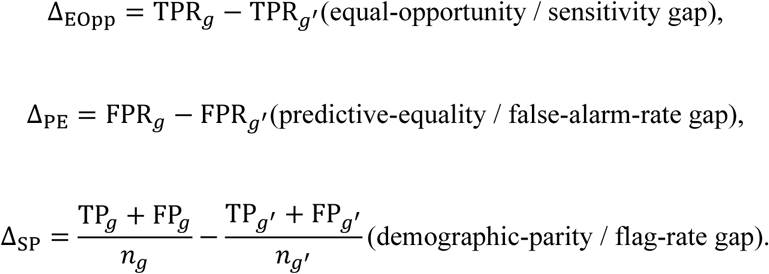

The equalized-odds gap is the larger of the two error-rate gaps, Δ_EO_ = max(|*Δ*_*EOpp*_| , | *Δ*_*PE*_|). For attributes with more than two levels, each gap is summarized by its largest absolute value over all pairs of levels.

#### Effect size

Each rate-based gap is reported with Cohen’s ℎ on the two underlying proportions *p*_*g*_, *p*_*g*_′,

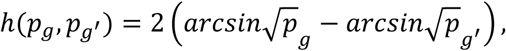

an effect-size magnitude less sensitive to the base rate than the raw difference.

### Calibration metrics

A model is calibrated within subgroup *g* if, among patients assigned predicted risk *p*, the observed infection rate is also *p*, i.e. *E*[ *Y* ∣ *p*, *A* = *g* ] = *p*. Calibration was assessed three ways.

i. **Calibration curves.** The conditional outcome probability *E*[*Y* ∣ *p*, *A* = *g* ] is estimated as a smooth function of *p* by LOESS, with 95% bootstrap bands. The identity line *y* = *p* is perfect calibration; a curve below the line indicates overestimation of risk.
ii. **Calibration intercept and slope** [34]. On the log-odds scale 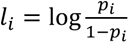, the intercept *α*_*g*_(calibration-in-the-large) is the constant of a one-parameter logistic regression on *I*_*g*_with the slope of *l*_*i*_ fixed to 1,

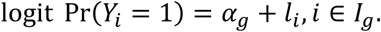

*α*_*g*_ = 0 is unbiased; *α*_*g*_ < 0 indicates systematic over-prediction (as observed globally here, *α* = −1.02). The slope *β*_*g*_ is the coefficient of *l*_*i*_ when both terms are free,

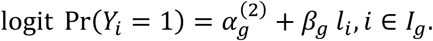

*β*_*g*_ = 1 means the predicted-risk gradient is correctly scaled in *g*; *β*_*g*_ < 1 means predictions are too spread out (over-extreme), *β*_*g*_ > 1 too compressed.

(iii) **Brier score, ECE, and ICI.** The Brier score is the mean squared distance between predicted probability and outcome,

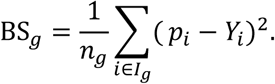

Using *K* = 10 equal-frequency bins, with bin *k* containing *n*_*g*,*k*_ predictions of mean predicted risk *p*_*g*,*k*_ and observed event rate *y*_*g*,*k*_, the expected calibration error is the prevalence-weighted mean absolute gap between predicted and observed risk,

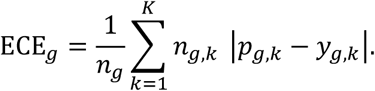

The integrated calibration index evaluates the same patient-by-patient gap against the LOESS curve *f*_*g*_ from (i),

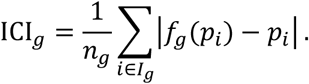

Hosmer–Lemeshow tests were not performed, given their well-documented sample-size sensitivity [39].

### Clinical-utility metrics

Decision curve analysis [38] was performed per subgroup across the threshold range *t* ∈ [0.02,0.40], the plausible band for intensified postoperative monitoring. The net benefit of a “flag and monitor when *p*_*i*_ ≥ *t*” policy is true positives gained minus false positives weighted by the harm ratio *t*/(1 − *t*) implied by the threshold itself,

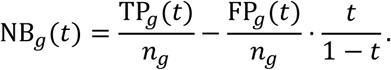

Because NB_*g*_(*t*) scales with the subgroup prevalence 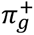, the standardized net benefit [23]

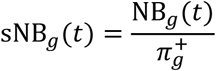

is also reported per subgroup, normalized so that sNB_*g*_(*t*) = 1 corresponds to a hypothetical perfect model on *I*_*g*_and sNB_*g*_(*t*) ≤ 0 to no benefit over the “treat-none” default. The primary clinical-utility gap statistic is the largest absolute net-benefit difference between any two subgroups, taken across the threshold band,

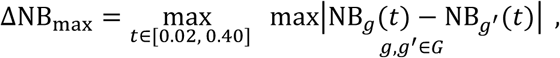

with bootstrap confidence intervals.

### Model-based interaction tests

Gap statistics describe what a fixed threshold sees on the calibrated probability scale; they do not, on their own, say whether a covariate’s effect on the outcome differs by sensitive attribute. To complement them, likelihood-ratio tests compare nested logistic regressions. For a covariate *X*, sensitive attribute *A* (sex), and adjustment vector *C* (age and ASA, as appropriate):

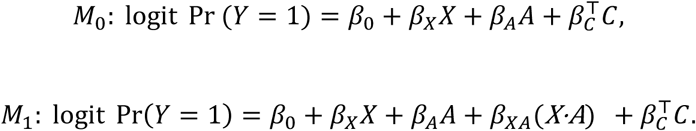

Under *H*_0_: *β*_*XA*_ = 0, the test statistic

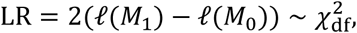

with df equal to the number of interaction parameters. Prespecified pairs were sex × physiological reserve and sex × ASA; a screen of sex × comorbidity interactions covered 23 comorbidities and was Benjamini–Hochberg–adjusted across that family. Each test is reported with its *p*-value and the Cox–Snell pseudo-*R*^2^ increment Δ*R*^2^, which guards against the common pattern of a small *p* but clinically negligible effect that a sufficiently powered cohort produces by default.

For the sex × comorbidity screen, the odds ratio for postoperative infection associated with each comorbidity was additionally estimated separately within each sex from its 2 × 2 table, with a Wald 95% confidence interval (Figure 5B; Supplementary Table 8). A within-sex odds ratio is an expected association and is not, by itself, evidence of a sex difference; the fairness-relevant comparison is the interaction term above.

### Bias-mitigation methods

Both mitigations are score-level postprocessors: they act on the model’s predicted probabilities and require no access to the training pipeline. To avoid evaluating mitigated probabilities in-sample, each was fitted by 5-fold stratified cross-validation (within every fold the transform was estimated on four folds (the calibration set 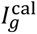) and applied to the held-out fold) so that every patient receives an out-of-fold mitigated prediction and the before-versus-after comparison is computed on the full cohort. Group-specific isotonic recalibration was applied independently to three axes (sex, physiological reserve, and procedure cluster); Wasserstein-barycenter postprocessing was applied to the procedure-cluster axis.

### Group-specific isotonic recalibration

For each subgroup *g*, a monotone non-decreasing step function *ι*_*g*_: [0,1] → [0,1] is fitted on the calibration set by minimizing the squared distance to the outcomes,

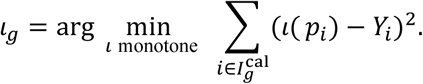

At test time the recalibrated probability for a patient in *g* is 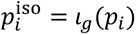. Because the map is monotone, the within-subgroup rank order, and hence within-subgroup AUROC, is preserved except for tie-induced rank collapses at very low event counts.

### Wasserstein-barycenter score postprocessing

Let *F*_*g*_ denote the empirical CDF of 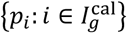. The one-dimensional Wasserstein barycenter, weighted by the group sizes *π*_*g*_, is the distribution whose quantile function is the weighted average of the per-group quantile functions,

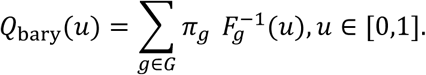

A prediction *p* from a patient in *g* is mapped to *p*^W^ = *Q*_bary_(*F*_*g*_(*p*)) and re-expressed as the value at its own within-group percentile, read off the common barycenter scale. The map is monotone within each subgroup, so within-group rank order and AUROC are preserved exactly; on the calibration set all per-subgroup score distributions coincide after the map, enforcing demographic parity at the score level. By the impossibility of jointly satisfying calibration and equalized odds when group base rates differ [44], score postprocessing was restricted to demographic parity, with equalized-odds and equal-opportunity gaps used only as evaluation diagnostics. Both outputs remain probabilities in [0,1] and so stay usable by the calibration and decision-curve analyses [41–43], in contrast to hard-threshold schemes whose output is a decision rather than a probability [32,33].

### Uncertainty quantification and multiple comparisons

#### Bootstrap confidence intervals

All confidence intervals are stratified bootstrap (fixed seed), with *B* = 200 resamples, drawn within subgroups and at the population level. Pairwise gap statistics reuse the same resampled distribution as their underlying within-group rates, so that the dependence between a gap and those rates is preserved.

#### Paired bootstrap

Between-group differences of overall metrics, e.g. ΔAUROC = AUROC_*g*_ − AUROC_*g*_′ uses a paired bootstrap that resamples patients from the pooled cohort and re-stratifies by group, preserving the dependence between the two subgroup estimates. Letting Δ_*b*_ denote the difference computed on resample *b*, the two-sided percentile *p*-value with continuity correction is

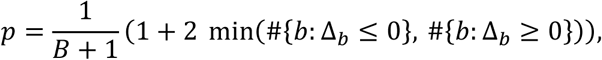

reported alongside the symmetric percentile interval [Δ_*α*/2_, Δ_1−*α*/2_] at *α* = 0.05.

#### Empirical-Bayes shrinkage

For the intersectional analysis, two-way cells with fewer than 50 outcome events have their cell rate *r*_*c*_ shrunk toward the marginal subgroup rate *r*_0_,

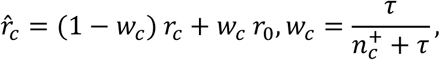

where 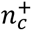 is the cell event count and *τ* is the shrinkage strength estimated by the method of moments from cross-cell variance. The pooling weight *w*_*c*_ (0 = no shrinkage, 1 = full shrinkage to the margin) is reported per cell (Supplementary Table 7) so that the degree of pooling is explicit.

#### Benjamini–Hochberg FDR

Multiple-comparison correction controls the false discovery rate at *q* = 0.05, applied independently within each of four test families: (a) the gap-statistical significance tests within each axis (discrimination, calibration, clinical utility); (b) the sex × comorbidity interaction screen; (c) the sex-stratified scan of cohort characterization variables (Table 1, Supplementary Table 1); and (d) the feature-by-sex effect-size scan (Figure 5A). Within a family of *m* tests, the ordered *p*-values *p*_(1)_ ≤ ⋯ ≤ *p*_(*m*)_ are compared with the linear thresholds (*k*/*m*) *q*; the largest *k*\*satisfying *p*_(*k*)_ ≤ (*k*/*m*) *q* is identified, and all hypotheses with *p* ≤ *p*_(*k*\*)_ are rejected. Within-subgroup point estimates are reported without correction.

### Effect sizes

The cohort characterization table and the per-subgroup gap annotations report an effect size matched to each comparison:

- **Cohen’s** *d* for a continuous variable compared across two groups: *d* = (*x̄*_1_ − *x̄*_2_)/*S*_*p*_, with *S*_*p*_ the pooled standard deviation.
- **Cohen’s** ℎ for a binary variable or proportion: ℎ = 2(arcsin√*p*_1_ − arcsin√*p*_2_).
- **Cramér’s** *V* for an unordered categorical variable in an *r* × *c* contingency table: 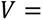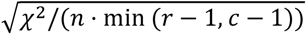
- *η*^2^ for a continuous variable stratified by an attribute with more than two levels: *η*^2^ = SS_between_/SS_total_.

Each effect size is accompanied by its conventional magnitude class (negligible / small / medium / large). Where a comparison is composite (the equalized-odds gap) or operates on a non-rate metric (calibration intercept or slope, decision-curve net benefit), no single effect size applies, and the column is left blank rather than filled with a misleading number.

### Outcome definition: ICD-10-GM code list

Bacterial postoperative infection within the index admission was defined by the presence of any code within the following 68 ICD-10-GM categories (280 codes), following the coding pipeline established for the Inselspital surgical cohort:

A02, A03, A04, A23, A26, A31, A32, A36–A42, A46, A48, A49, A54–A56, B95, B96, B98, G00, G01, G04, G05, H44, H60, H62, H66, I32, I41, J02, J03, J13–J17, J20, J86, K12, K35, K63, K65, K67, L00, L01, L03, L08, L51, M00, M01, M46, M63, M68, M72, M73, M86, N13, N49, N74, N76, U69, U80, U81, U82.

Where present-on-admission (POA) flags were available, infections coded as already present at admission were excluded, removing false positives of non-postoperative origin. The model treats infection as binary, and this audit follows the same convention.

### SUPPORT2 cohort

As a portability check, the identical audit was applied to the public SUPPORT2 cohort (Study to Understand Prognoses and Preferences for Outcomes and Risks of Treatment; UCI Machine Learning Repository [45]), comprising 9,105 critically ill adults from five US medical centers. On this cohort the audited model is a logistic-regression classifier for in-hospital mortality, fitted on a training partition and audited on a held-out test partition (*n* = 1,821; mortality 25.9%). All metric definitions apply unchanged. The SUPPORT2 audit additionally exercises a race/ethnicity axis (sex and age stratum being shared with the Bern audit), which the Swiss EHR does not record, and was run end-to-end from the example script shipped with the *isitfair* package under a fixed random seed. Group-specific isotonic recalibration was applied to the race/ethnicity axis. This secondary cohort is reported only to demonstrate portability and is not pooled with the analysis from our cohort.

## Notes

### Competing Interest Statement

The authors have declared no competing interest.

### Author Declarations

The study was approved by the Cantonal Ethics Committee of Bern (KEK-ID: 2021-00965), with a waiver of informed consent due to the study retrospective design.

